# What tools do men need to make an informed decision about germline genetic testing for prostate cancer? A qualitative and survey study

**DOI:** 10.64898/2026.03.27.26349466

**Authors:** Kelsie Raspin, Larissa Bartlett, Jennifer K Makin, Rebecca Wilson, Kate Butorac, Liesel M FitzGerald, Jessica Roydhouse, Joanne L Dickinson

**Author notes:** These authors contributed equally to this work (Jessica Roydhouse, Joanne L Dickinson).

## Abstract

**BACKGROUND:** Germline genetic testing for prostate cancer (PrCa) can help identify those at greater risk of developing advanced or lethal disease, while also influencing treatment selection, however it is not widely embedded in PrCa care. An integral part of developing frameworks for the delivery of precision medicine is the inclusion of patients/consumers in the design to ensure their needs are met.

**METHODS:** *Phase I*: Focus group discussions with men diagnosed with PrCa (n=20), to determine their opinions, perceptions and expectations of germline genetic testing for PrCa. Data were analysed thematically. *Phase II: Phase I* themes informed a *Precision Medicine in Prostate Cancer Information Toolkit*, which was reviewed by patients (n=14), their carers/family members (n=4) and healthcare providers (n=14).

**RESULTS:** *Phase I*: Knowledge about precision medicine and genetic testing was generally low. The strongest motivation for undertaking testing for patients was to determine family members’ risk (n=7), and the biggest concern pertained to insurance discrimination (n=5). *Phase II:* Healthcare providers (n=8) generally found the purpose of the toolkit to be clearer than patients (n=5), who found it difficult to imagine its usefulness when considering their own genetic risk at/before diagnosis. Information regarding life insurance, implications for their family and costs associated with testing were of concern to all participant groups.

**CONCLUSIONS:** This study revealed critical knowledge gaps, preferred communication/support needs, and concerns/risks associated with germline genetic testing for PrCa. Our toolkit, whilst valuable, requires further refinement to ensure broad accessibility across literary and cultural contexts.

## 1. INTRODUCTION

Prostate cancer (PrCa) is the most common non-cutaneous cancer worldwide, and among Australian men, the mortality rate is second only to that of lung cancer ^1^. PrCa has the highest heritability of all the common cancers ^2^, yet the integration of germline genetic testing into PrCa clinical care has significantly lagged behind that of other cancers, particularly breast and ovarian^3^. Germline genetic knowledge, identified using genetic testing, can identify those likely to develop life threatening disease, determine the most appropriate therapy and inform early screening of family members.

Australia’s healthcare system is reliant on a publicly funded subsidised arrangement that covers the diagnosis and treatment of PrCa. However, the only subsidised germline genetic test for PrCa is for patients with metastatic castration-resistant disease where tumour testing is not clinically feasible (Medicare Benefits Schedule – Item 73304). These patients are tested for germline variants in *BRCA1/2* to determine whether they are eligible for treatment with Olaparib, a targeted therapy. Access to this genetic test is not equitable across Australia ^4^, with regional, rural and remote communities often under-resourced to provide such services within a useful timeframe. Notably, over 28% of Australia’s population is living in regional/rural/remote areas and these regions experience significant disparities in outcomes ^5^.

Equitable access to genomic PrCa medicine is critical for regional, rural and remote areas across the globe, given the significantly poorer outcomes for PrCa patients in these communities. Men from these areas access diagnostic and treatment services less frequently ^6,7^, face longer delays between diagnosis and treatment initiation ^8,9^, and significantly, are diagnosed later ^8,10^ and with more advanced, higher-risk disease ^8,10–12^. They are also less likely to seek genetic services ^4^ and have limited opportunities to participate in clinical trials or benefit from precision medicine ^13,14^. Such disparities are likely due to a combination of differences in socioeconomic and educational status, race/ethnicity, distance to health services and access to health expertise required to deliver health innovations ^14–19^. Our previous research has shown that even within a wholly regional state, there are still varying levels of disparities ^8^. To reduce these disparities, it is important to understand what barriers, particularly educational/resource/support barriers, are preventing men with PrCa from accessing these genetic services.

An integral part of healthcare delivery design is engagement with patients to develop appropriate information sharing tools and to provide them with the opportunity to shape healthcare delivery catered to their needs ^20,21^. Studies have shown that patients’ knowledge, understanding and attitudes towards precision medicine play a critical role in determining how effectively these approaches can be implemented to improve health outcomes ^15,22^, though very few studies have co-designed tools for precision medicine ^23^. Most notably, to the best of our knowledge, no prior studies have published a co-designed PrCa-specific precision medicine tool with patients. Thus, this two-phase study aimed to address this gap by working with people affected by PrCa to develop and evaluate an information toolkit to support patients undergoing germline genetic testing for PrCa.

## 2. METHODS

Informed consent was obtained from all participants included in this study. This study was approved by the University of Tasmania’s Human Research Ethics Committee (H0026835).

### 2.1 Phase I

In *Phase I*, we engaged with PrCa patients/consumers to determine their opinions, perceptions and expectations of genetic testing in the context of PrCa. Through the team’s networks, including local community groups (Cancer Council Tasmania & Hobart Probus Club) and participating clinical healthcare providers (Urologists/Oncologists), we invited men diagnosed with PrCa to participate in focus groups held between December 2022 and September 2023. Focus groups were selected because they are well suited to eliciting participants’ attitudes and lived experiences of disease, with group interaction facilitating the exploration of diverse perspectives ^24^. Participants were advised that groups may take up to 90 min. Group sizes were capped at 4-8 participants to ensure representation of all group members. For each of the four focus groups, one facilitator (JR or KR) and a support person for note taking (RW or KB) were present from the investigator team. The first focus group was facilitated by JR, a female researcher with experience in qualitative and quantitative methods, with KR, a female researcher with experience in genetics research, as the support person and note taker. JR is experienced in conducting focus groups with people affected by cancer and provided training and ongoing support to KR to facilitate the other three. RW, a female with experience in working with people affected by cancer in both a research and clinical setting and KB, a female registered nurse with decades of experience caring for people affected by cancer, supported the remaining groups, including note taking.

Focus group discussions were guided by a list of open-ended questions (Supplementary Information 1) that focussed on patients’ concerns and expectations regarding the use of genetic information in the diagnosis and treatment of PrCa, as well as their information and support needs, including collection of patient health status and other such information through questionnaires. Focus groups were tape-recorded and the recordings transcribed verbatim by an external transcribing company. One focus group was not recorded due to a technical issue, but detailed notes were taken by the research team, and these were included in the analysis. Transcripts were de-identified by KR for analysis.

#### Analysis

Coding of the de-identified transcripts began once two focus groups were completed, with additional transcripts coded when available. No further focus groups were conducted once saturation was achieved (i.e., there were only minimal new themes emerging from the analysis)^25^. De-identified transcripts and notes were imported into the qualitative data management program NVivo 12. Data were analysed thematically by an experienced qualitative researcher (JM) in line with the questions that guided the focus groups. Detailed sub-theme codes were used to capture the broadest range of themes possible to inform *Phase II*. Emerging themes were reviewed by a second member of the research team (JR).

### 2.2 Phase II

The themes and sub-themes identified in *Phase I* were used to design and develop a *Precision Medicine in Prostate Cancer Information Toolkit*. In *Phase II*, the toolkit was evaluated by patients, carers/family members, and healthcare providers to assess its clarity, understandability, and readability to inform a second iteration. Carers/family members were invited to participate through the patients themselves and are unidentifiable. We assume that these participants are spouses or biological family members.

The toolkit was initially developed by KR and further edited by JR and JLD. Participants were then invited to review the toolkit prior to participating in a focus group discussion or survey. Patients were asked to participate in two focus groups held in June and July 2024. Focus group discussions were guided by a list of open-ended questions (Supplementary Information 2), with discussions tape-recorded and the recordings transcribed verbatim by an external transcribing company. Due to limited availability and scheduling difficulties, only patients attended the focus groups, while carers/family members and healthcare providers were invited to complete an e-survey through RedCap (Research Electronic Data Capture database) ^26,27^, which included the same questions asked in the focus groups. Survey responses were then exported from RedCap for analysis in Excel.

#### Analysis

Emergent themes in the focus group transcript data were identified using *Leximancer* ^28^, and the survey responses were manually coded in Excel. This round of analyses was conducted by an independent researcher, LB, with expertise in psychometric and qualitative data handling. The commonly identified themes around the toolkit’s acceptability and readability were highlighted and compared across both sets of results. Themes were reviewed by two other members of the research team (KR and JR).

## 3. RESULTS

### 3.1 Phase I

A total of 20 men diagnosed with PrCa, aged between 50-83 (average=72), participated in four focus groups across *Phase I*. Their PrCa treatment management included active surveillance (n=1), radical prostatectomy (n=11), radiation (n=10) and hormone therapy (n=5). Many men experienced severe treatment side-effects, while six men experienced PrCa recurrence or progression to metastatic disease. Most participants (n=14 of 20) underwent more than one treatment type and data were missing for some participants. Of the 20 participants, three had a previous experience of genetic testing in relation to their PrCa. All *Phase I* focus group themes are presented in Supplementary Table 1 and described in detail below.

#### Understanding of the term ‘Precision Medicine’

In *Phase I*, nine of 20 participants had heard of precision medicine, but did not necessarily understand its meaning (Table 1).

> *…. I have no understanding of it. I’ve heard the term, but I don’t know what it means.* (P10)

**Table 1.** *Phase I* Themes – understanding and attitudes of precision medicine.

| Name/Description | Files<sup>1</sup> | References<sup>2</sup> |
| --- | --- | --- |
| **Precision Medicine** | **5** | **32** |
| *Understanding* | 5 | 26 |
| a) Avoiding unnecessary treatment and side effects | 1 | 1 |
| b) Don't understand | 4 | 9 |
| c) More effective treatment | 1 | 1 |
| d) More precise or targeted medicine | 2 | 4 |
| e) Targeted to cancer | 1 | 1 |
| f) Targeted to individual | 3 | 7 |
| g) Therapy focused on one location | 2 | 3 |
| *Attitudes* | 2 | 6 |
| a) Equity of access and cost | 1 | 1 |
| b) Pro | 2 | 5 |
| c) Pro - if outcome is better | 1 | 1 |
| d) Pro - innovation is good | 1 | 2 |
| <sup>1</sup>Number of focus group transcripts. Noting that three focus group transcripts were included, in addition to two team members notes for the fourth group that was not recorded due to technical difficulties. Therefore, n=5.<br><sup>2</sup>Number of participants in total that referred to the specific theme. |  |  |

Of those that did understand the term, many were correct in its definition, and two men thought that it was more effective.

> *…very targeted, precise, for your own needs, that’s what I would say it was.* (P1)
>
> *…. If you can understand the gene sequence of the cancer cell, then you can, I believe, probably find a drug that will target that specific cell more without all the side effects. I think.* (P19)
>
> *…. I think it’s about not putting people through unnecessary treatments because treatments come with a lot of side effects.* (P7)

Two men who had never heard of the term could guess the meaning based on the use of ‘precision’, including one individual in a focus group where none of the participants had heard of the term (n=4) (Table 1).

#### Attitudes towards Precision Medicine

Whilst all groups were prompted, only two focus groups deeply explored participants’ attitudes towards precision medicine (Table 1). Seven of the ten participants were positive and receptive.

> *…. It sounds good to me.* (1)
>
> *…. I think if it provides a better outcome, I think that’s the crucial part of it and just the terminology, ‘precision’ would give you hope that that’s what it would do.* (P2)

One man noted that there will obviously be equity issues pertaining to precision medicine.

> *…. There’ll be - inevitably, there’ll be issues around equity of access and cost, particularly initially.* (P4)

#### Understanding of the term ‘Genetic Testing’

All 20 participants had heard of genetic testing, but not specifically genetic testing for PrCa. Some participants had a general understanding of what genetic testing was (Table 2).

> *…to me, it’s about testing people’s genetic profiles for mutations that might lead to particular forms of cancer.* (P7)

**Table 2.** *Phase I* Themes – understanding and attitudes of genetic testing.

| Name/Description | Files | References |
| --- | --- | --- |
| **Genetic Testing** | **5** | **191** |
| *Understanding* | 4 | 20 |
| a) Unsure or need more information | 4 | 8 |
| b) Know familial risk | 3 | 6 |
| c) Way of finding best treatment | 3 | 4 |
| d) Mutations associated with cancer | 1 | 2 |
| e) Would like to know how genes are associated | 1 | 2 |
| f) Currently no treatment following on | 1 | 1 |
| g) Unsure about additional benefits compared with current testing | 1 | 1 |
| *Attitudes* | 4 | 9 |
| a) Pro | 2 | 3 |
| b) Understanding familial risk would be helpful | 3 | 5 |
| c) No complaints about usual care treatment (without testing) | 1 | 1 |
| <sup>1</sup>Number of focus group transcripts. Noting that three focus group transcripts were included, in addition to two team members notes for the fourth group that was not recorded due to technical difficulties. Therefore, n=5.<br><sup>2</sup>Number of participants in total that referred to the specific theme. |  |  |

However, eight participants indicated that they were unsure of what genetic testing showed.

> *…. I’m not completely clear but I understand from the various forums I’ve been in that it’s a new way of finding the best treatment for cancer using a method which is non-invasive but certainly quite new.* (P5)

Six of 20 participants understood that genetic testing can provide you with information about risk to self and family members. Other reported understandings of genetic testing included identifying mutations associated with cancer and informing the choice of treatment, which was presumed to lead to better outcomes. Two participants thought that genetic testing could provide them with information about the aggressiveness of their cancer (Table 2).

#### Attitudes towards Genetic Testing

In terms of their attitudes (Table 2), 40% of the participants were in favour of genetic testing for their PrCa (8 of 20). Their main reason was believing that understanding familial cancer risk would be helpful (5 of 8 participants).

> *… Currently, it would be of help to help my brother and his family because for me, I don’t have a son to pass it on to.* (P5)

One man noted that he had no complaints about his care, which did not include genetic testing.

> *… I have to say that I’ve got absolutely no complaints about any of my treatment. I’ve found that all of the people that I’ve seen, the specialists have been very attentive, very interested, very flexible and decent human beings.* (P12)

#### Benefits of Genetic Testing

The most frequently mentioned reasons to do genetic testing included, 1) inform family members’ risk (n=7), 2) help identify most effective treatment options (n=6), 3) lead to early detection and prevention (n=5), and 4) reduce use of less effective or more onerous therapies or tests (n=3) (Table 3).

> *…. I think, for my son, if he was aware that he had the markers of an abnormal prostate, then it would flag him to be monitored more closely than to live life blindly without-and then finding that out too late [when the cancer has spread].* (P19)

**Table 3.**
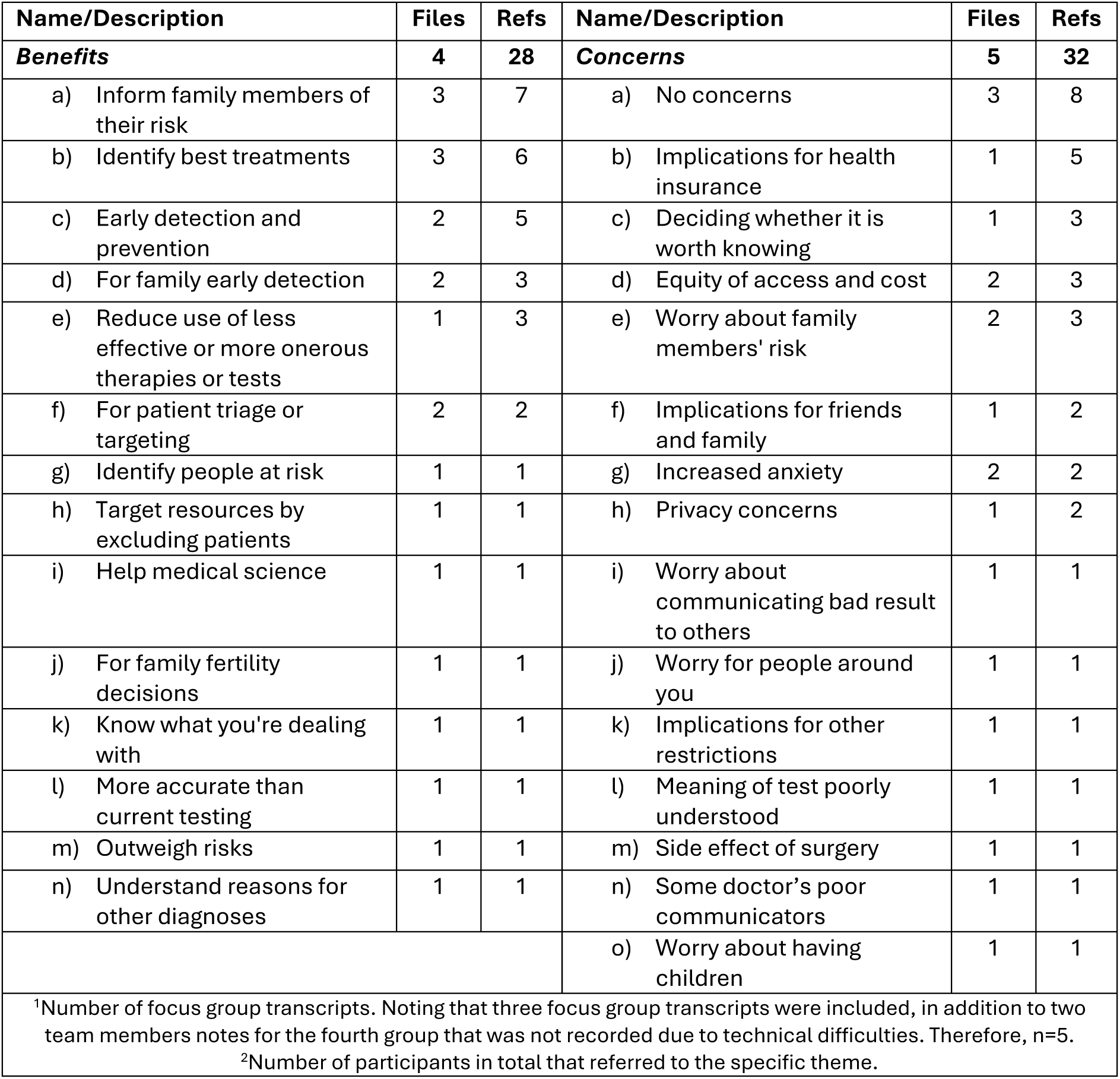
*Phase I* Themes – perceived benefits and concerns of genetic testing.

| Name/Description | Files | Refs | Name/Description | Files | Refs |
| --- | --- | --- | --- | --- | --- |
| **Benefits** | **4** | **28** | **Concerns** | **5** | **32** |
| a) Inform family members of their risk | 3 | 7 | a) No concerns | 3 | 8 |
| b) Identify best treatments | 3 | 6 | b) Implications for health insurance | 1 | 5 |
| c) Early detection and prevention | 2 | 5 | c) Deciding whether it is worth knowing | 1 | 3 |
| d) For family early detection | 2 | 3 | d) Equity of access and cost | 2 | 3 |
| e) Reduce use of less effective or more onerous therapies or tests | 1 | 3 | e) Worry about family members' risk | 2 | 3 |
| f) For patient triage or targeting | 2 | 2 | f) Implications for friends and family | 1 | 2 |
| g) Identify people at risk | 1 | 1 | g) Increased anxiety | 2 | 2 |
| h) Target resources by excluding patients | 1 | 1 | h) Privacy concerns | 1 | 2 |
| i) Help medical science | 1 | 1 | i) Worry about communicating bad result to others | 1 | 1 |
| j) For family fertility decisions | 1 | 1 | j) Worry for people around you | 1 | 1 |
| k) Know what you're dealing with | 1 | 1 | k) Implications for other restrictions | 1 | 1 |
| l) More accurate than current testing | 1 | 1 | l) Meaning of test poorly understood | 1 | 1 |
| m) Outweigh risks | 1 | 1 | m) Side effect of surgery | 1 | 1 |
| n) Understand reasons for other diagnoses | 1 | 1 | n) Some doctor's poor communicators | 1 | 1 |
|  |  |  | o) Worry about having children | 1 | 1 |
| <sup>1</sup>Number of focus group transcripts. Noting that three focus group transcripts were included, in addition to two team members notes for the fourth group that was not recorded due to technical difficulties. Therefore, n=5.<br><sup>2</sup>Number of participants in total that referred to the specific theme. |  |  |  |  |  |

Less commonly mentioned benefits included, 1) assist with patient/resource triage (n=1), 2) help medical science (n=1), and 3) more accurate than current testing (n=1).

> *…it would be a kind of sifting process I guess about where you would put extra resources.* (P10)

Additional perceived benefits of genetic testing for PrCa are displayed in Table 3.

#### Concerns of Genetic Testing

Sixty percent of the participants (12 of 20) had concerns pertaining to genetic testing for their cancer (Table 3). Of the concerns raised, the most common included, 1) compromised ability to get health/life insurance (n=5), 2) anxiety associated with deciding whether to undergo genetic testing (n=3), and 3) equity of access and cost (n=3).

> *…huge benefits of understanding all of this but then that could definitely elicit a lot of anxiety that you need to manage somehow…* (P4)

In terms of insurance, two participants directly referred to ‘health insurance’, whereas other participants spoke of insurance more broadly. However, further discussion within each focus group indicated that most were referring to life insurance.

> *…. I do have one concern, the implications to health insurance.*(P1)
>
> *…. I think the insurance scenario is real. If I had got the wrong answer with the testing to me in relation to what genes my daughters might’ve inherited, there might be a point where they’re seeking insurance, they have to disclose that and the answer is, “No, you can’t have insurance.”* (P7)

Less common concerns included, 1) worry about communicating the results (n=1), 2) privacy concerns (n=2), and 3) worry pertaining to variants of uncertain significance (n=1). A full list can be found in Table 3.

> *…. “There was worry about if you get the wrong answer [from the genetic test], how are you going to handle that in conversation and informing these people and when are you going to do it, because there are no absolute guarantees, it’s about risk.”*(P7)

#### Desired Information/Source/Support for Genetic Testing

Participants mainly wanted simplified information about, 1) method of testing, i.e., what is involved (n=3), 2) how will their result help decide which treatment is best (n=3), 3) benefit (n=2), 4) cost (n=2), and 5) data risks (n=2) (Table 4).

> *…. Cost, benefit, outcomes and I guess risks in terms of your data and anxiety and things like that.* (P4)
>
> *…. And you need to make it simplistic because it’s going to blokes and blokes will just - won’t read it…* (P3)

**Table 4.** *Phase I* Themes – desired information to undergo genetic testing.

| Name/Description | Files | References |
| --- | --- | --- |
| **Desired information** | **4** | **36** |
| a) How will it help decide treatment | 2 | 3 |
| b) Method and experience of testing | 2 | 3 |
| c) Benefit | 1 | 2 |
| d) Cost | 2 | 2 |
| e) Data risks | 1 | 2 |
| f) No questions | 1 | 2 |
| g) Outcomes | 1 | 2 |
| h) Requires understanding of basic risk concepts | 2 | 2 |
| i) Timing - for prevention or treatment guidance | 1 | 2 |
| j) What information does the test give | 1 | 2 |
| k) What prompts testing | 1 | 2 |
| l) What questions to ask | 1 | 2 |
| m) Benefit for choosing treatment | 1 | 1 |
| n) Best testing model | 1 | 1 |
| o) How information is used and collected | 1 | 1 |
| p) How reliable is result | 1 | 1 |
| q) If tested, can family members access result | 1 | 1 |
| r) Is result indicator of risk or actual tumour | 1 | 1 |
| s) Other considerations in deciding whether to test | 1 | 1 |
| t) Risks - anxiety | 1 | 1 |
| u) Socioeconomic equity | 1 | 1 |
| v) Timing of results | 1 | 1 |
| w) What do the results mean | 1 | 1 |
| <sup>1</sup>Number of focus group transcripts. Noting that three focus group transcripts were included, in addition to two team members notes for the fourth group that was not recorded due to technical difficulties. Therefore, n=5. |  |  |
| <sup>2</sup>Number of participants in total that referred to the specific theme. |  |  |

Many participants also said that “*you don’t know what you don’t know”*.

In terms of who should provide this information, seven out of the 20 participants would like to receive this information from their medical practitioner and some highlighted that they would need some sort of counselling.

> *…. Probably more the genetic counsellor, who’s explaining what’s going on and would go through what they discovered…* (P19)

However, many said that they would be happy to speak to the doctor/specialist who ordered the test to obtain information regarding the meaning of the result. Two participants mentioned that they would want to be supported by a male prostate cancer nurse, in addition to a genetic counsellor.

> *…. A support person, a male nurse.*(P17)

One participant highlighted that information would ideally be provided through a specialised genetics or cancer clinic.

#### Questionnaires pertaining to Genetic Testing

All 20 participants agreed that they would feel comfortable answering any questions relating to their health, given their own PrCa experience. However, they referred to there being a lack of privacy when you are a cancer patient (n=5). One participant suggested that they would prefer to be interviewed rather than answer a questionnaire, whereas another stated that intimate information can be easier in written form.

> *…. Questionnaires aren’t as nice as sitting at the table and talking. I’d be happy to tell you something that maybe I wouldn’t want to write down.* (P20)
>
> *…. Depends on how intimate the information is. Sometimes it’s more impersonal on a questionnaire and you can feel more free to relay that information.* (P19)

There were two references to the questionnaire including closed rather than open questions due to the assumed low understanding of respondents.

> *…. Yes, you couldn’t just give us a questionnaire and say, ‘Write down what you’re concerned about’, because we don’t know what there is to be concerned about.* (P20)

### 3.2 Phase II

In *Phase II*, a *Precision Medicine in Prostate Cancer Information Toolkit* was designed and developed using the *Phase I* data (available upon request). The toolkit was then reviewed by 14 men with PrCa, all of whom participated in *Phase I*, as well as 4 carers/family members and 14 healthcare providers, including medical oncologists (n=3), radiation oncologists (n=2), a urologist (n=1), a haematologist (n=1), prostate cancer specialist nurses (n=3), geneticists/clinical genetic counsellors (n=2) and pathologists (n=2).

When questioned whether the toolkit’s purpose was clear, five of 14 patients (36%), eight of 14 healthcare providers (57%) and all four carers/family members (100%) felt the purpose was clear, nine patients and five healthcare providers suggested that it could be improved, and one healthcare provider thought that the purpose was not clear at all (Table 5).

> *…. I might have a little bit of trouble I think. I don’t think it is all that clear at the outset.* (P3)
>
> *…I think there need to be more clarity re somatic and germline testing - indications, timing, rationale etc.* (HP1)
>
> *…. It may be clearer if there is an opening statement e.g. after viewing the toolkit you should understand…and a bit of a summary at the end however overall it is clear.* (HP2)
>
> *….Yes a really easy read format that went step by step.* (C/FM4)

**Table 5.** *Phase II* – purpose and usefulness of the toolkit.

|  | **Response** | **Patients**<br>**(n=14)** | **Healthcare**<br>**Providers**<br>**(n=14)** | **Carers/Family**<br>**Members**<br>**(n=4)** |
| --- | --- | --- | --- | --- |
| **Is the purpose clear?** |  |  |  |  |
|  | no | 0 | 1 | 0 |
|  | could be improved | 9 | 5 | 1 |
|  | yes | 5 | 8 | 3 |
| **Do you think others/patients will understand how things are described?** |  |  |  |  |
|  | no | 0 | 0 | 0 |
|  | could be improved | 7 | 6 | 0 |
|  | yes | 2 | 8 | 4 |
| **Are medical/scientific terms well defined?** |  |  |  |  |
|  | no | 0 | 0 | 0 |
|  | could be improved | 2 | 4 | 1 |
|  | yes | 7 | 10 | 3 |
| **Would the Toolkit be helpful if you/your patient were offered genetic testing?** |  |  |  |  |
|  | no | 0 | 0 | 0 |
|  | could be improved | 8 | 4 | 0 |
|  | yes | 5 | 10 | 4 |

Two patients, eight healthcare providers and all four carers/family members felt that other patients/their patients would understand the content of the toolkit, with the remaining feeling as though the content could be improved (Table 5).

> *…. I found because we’ve lived it I found the toolkit was quite easy to go through, but then I thought, hang on, what about a person who’s just about to start their journey or who hasn’t lived it before and doesn’t have all this information.* (P5)
>
> *…. Yes, the tool kit had little medical jargon and would suit patients with low health literacy.* (HP5)
>
> *…. At times, vocabulary may be too complex for some patients, however explanations are helpful.* (HP10)

Notably, only 5 patients (36%) thought the toolkit would be helpful if they were offered genetic testing, whereas 10 healthcare providers (71%) and all four carers/family members (100%) thought it would be helpful for others/their patients undergoing genetic testing for PrCa (Table 5).

As for medical terms, 11 of 14 patients thought they were well defined compared to 10 of 14 healthcare providers and 3 of 4 carers/family members (Table 5).

> *…. I found the language wasn’t too technical. Being a nonmedical person, I understood it reasonably well…* (P1)
>
> *…. Some words, both scientific and otherwise, may be too complex, i.e. prognosis, intervene, characteristics, conventional, adverse event, inherited.* (HP10)

The most common terms used in the toolkit that were not understood by patients and carers/family members were ‘panel’ (n=7) and ‘PARP’ (Poly (ADP-ribose) polymerase) (n=2), whereas there weren’t any terms that the healthcare providers didn’t understand.

> *…the one thing I stumbled on… was they’re talking about a panel. I didn’t get that. What panel?* (P4)
>
> *…. I just found PARP, that sounds really interesting, but then it was like … what is it?* (P6)
>
> *…. The only confusion was a dot point referred to a “panel” but I didn’t recall seeing that term defined or explained earlier in the tool kit.* (C/FM4)

In terms of missing information, patients highlighted missing details on 1) cost, 2) risk, 3) availability of test, 4) PARP eligibility and definition, 5) insurance, 6) timing in the cancer journey, and 7) personal stories of patients (Table 6). Healthcare providers thought that consent/privacy was not addressed, as well as test procedures, possible rebates available and the genes included on the panel (Table 6). Two carers/family members highlighted missing details on cost and how many men with PrCa are likely to benefit from precision medicine.

> *…. It would boil down to me for how much it costs and what are the insurance impacts. Do you get any on private health and things like that.* (P8)
>
> *…. Yeah, more case studies, exactly. Someone who’s got aggressive cancer or someone that’s got less aggressive or maybe watch and wait.* (P11)
>
> *…. The discussion around life insurance is too brief.* (HP2)
>
> *…. Different type of genetic testing specific to prostate cancer and timing of these tests and ramifications some test MBS [government-funded] rebatable…* (HP1)
>
> *…. I would add information about how the information is stored / used following genetic testing e.g. comment about privacy.* (HP2)

**Table 6.**
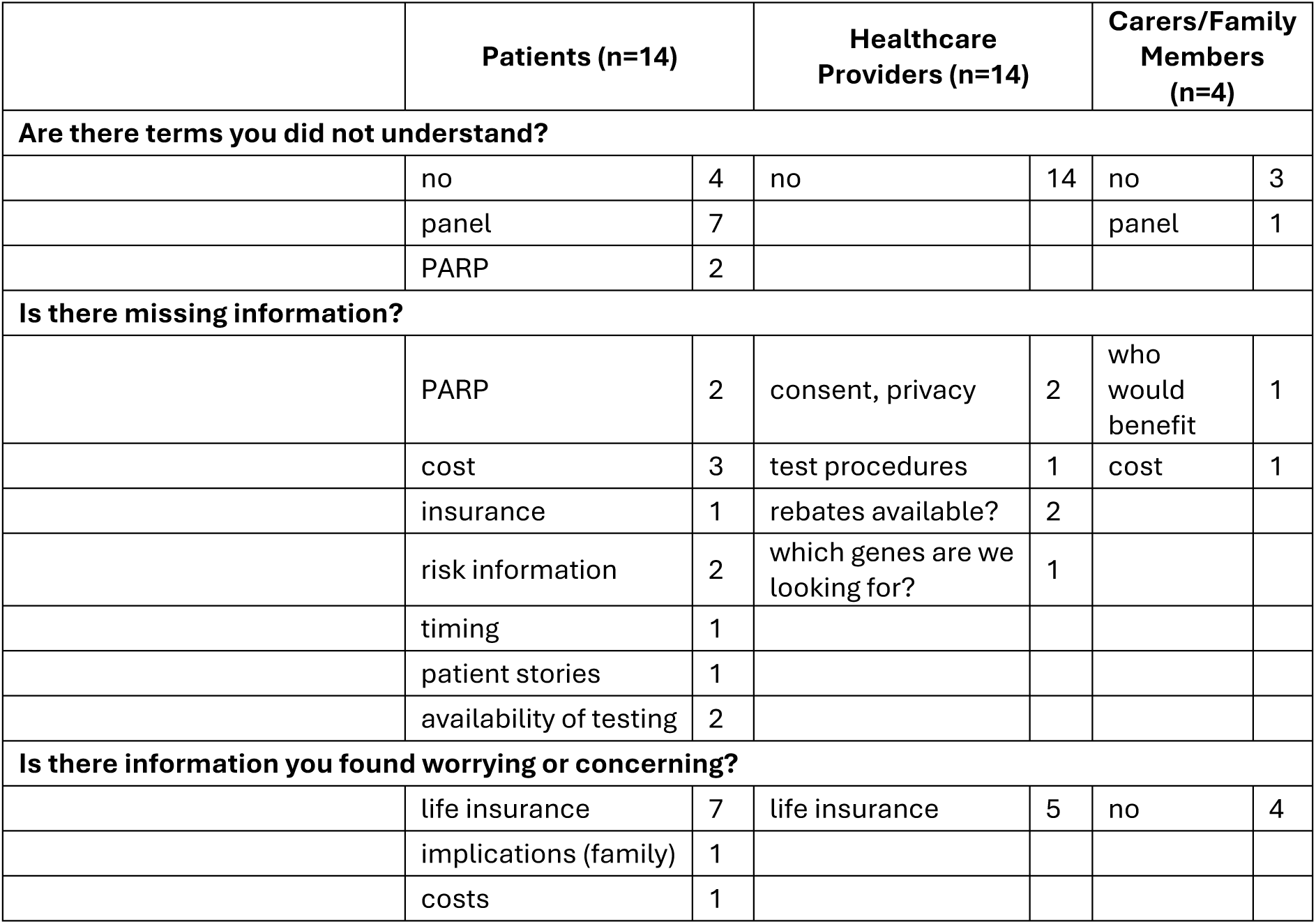
*Phase II* – missing and worrying information in the toolkit.

Both patients (7 of 14, 50%) and healthcare providers (5 of 14, 36%) found the information regarding life insurance worrying (Table 6), whereas none of the carers/family members did. Two patients also mentioned that they found the potential implications for their family (n=1) and the costs associated with undergoing genetic testing (n=1) worrying (Table 6).

> *…. Possible to think that some people might be quite concerned about the fact that they do have a genetic variation that might affect their family further on.* (P14)
>
> *…. Perhaps the implications for life insurance. Patients need to know that premiums can be loaded if they even have the test, irrespective of what it shows.* (HP9)

## 4. DISCUSSION

This study examined men’s understanding, attitudes and informational/support needs regarding germline genetic testing for PrCa within the context of precision medicine. Our *Phase I* findings revealed critical gaps in awareness, perceived benefits, and barriers associated with germline genetic testing for PrCa in a regional area amongst men with PrCa. Despite this, our study highlighted strong receptivity to genetic testing among PrCa patients, with family benefit the primary motivator for testing. Our patients also emphasised the need for clear and trusted information, in addition to PrCa and gender-specific support roles during the genetic testing process. In *Phase II*, evaluation of the co-designed *Precision Medicine in Prostate Cancer Information Toolkit* revealed that over a third of men understood the purpose of the toolkit and most did not find the language too technical, which contrasted with the views of carers/family members and healthcare providers. This highlights the need for multiple rounds of co-design with patients to ensure that the information provided is appropriately framed around patients lived experiences of cancer and genetic testing, including real patient stories to enable personal resonance and emotional relevance throughout the genetic testing pathway. It also needs to clearly articulate the benefits/limitations of genetic testing, as well as patient concerns pertaining to genetic discrimination, privacy and insurance discrimination.

Nearly half of our study’s participants (45%) were aware of the term precision medicine, although comprehension was limited, which is consistent with the literature – the concept remains poorly understood by patients in general (reviewed in Ahmed *et al.,* 2023 ^29^). Similarly, Stallings and colleagues (2023) ^30^ found that 18 of their 26 (69%) American English-speaking cancer patients/carers/family members reported little familiarity with the term precision medicine. Eleven believed genetics had little impact on their health, though their type of cancer was not reported ^30^. Furthermore, while all our participants were familiar with genetic testing more generally, few understood its application to PrCa, but all were receptive about undertaking genetic testing. It is worth noting that most published studies measuring patient knowledge, understanding and awareness of precision medicine have been conducted in the context of tumour or somatic testing (as opposed to germline testing) in breast, lung and colorectal cancer. Whilst we did not assess this in our study, a multinational survey of 811 patients and 895 physicians by Ciardiello and colleagues (2016) ^31^ found differences between perceived and actual understanding of tumour testing, where 78% of patients thought they understood, but only 23% of physicians agreed that their patients were fully informed. Conversely, in a recent study of 113 cancer patients (44% breast, 27% colon, 9% lung), as few as 28% felt they had enough knowledge to make informed decisions about undergoing genetic testing for their tumour to inform clinical management ^32^. These knowledge gaps underscore the need for targeted educational strategies, including tools and resources, that clearly articulate the purpose, process, and implications of genetic testing in cancer care, but particularly in the context of PrCa where research is limited – a key of objective of our study.

Patients identified several perceived benefits of genetic testing, with informing family members’ risk the most common motivator, followed by informing treatment decisions, as well as early detection and prevention for unaffected men. Our main finding reflects strong intergenerational concern among men with PrCa and aligns with the National Comprehensive Cancer Network’s guidelines emphasising the role of genetic testing in guiding familial risk management ^33^. Likewise, 33 PrCa patients in a qualitative study ^34^ also strongly valued testing for family benefit/cascade testing and personal utility to help with their treatment decisions.

While many of our patients reported no concerns with genetic testing, the concerns expressed were significant and warrant attention. Insurance discrimination was the most frequently cited worry, consistent with broader evidence that insurance-related barriers remain a major obstacle to genetic testing uptake. Concerningly, many of our participants used the terms ‘health’ and ‘life’ insurance interchangeably, although, in Australia, health insurers cannot refuse health insurance based on a genetic testing result. Australian research consistently shows low insurance literacy ^35,36^, with evidence suggesting that many misunderstand the purpose of life insurance and confuse it with health-related coverage. This misinterpretation isn’t specific to Australia, with many American studies reporting similar findings ^37^. Regarding life insurance, at the time of these focus groups, there was a moratorium in place by the then Australian peak insurance body, the Financial Services Council (FSC Standard 11: Moratorium on Genetic Tests in Life Insurance) on the use of genetic testing in life insurance policies, although this was limited. However, on April 8^th^ 2026, the Treasury Laws Amendment (Genetic Testing Protections in Life Insurance and Other Measures) Act 2026 (the Act) was enacted, which prevents the use of genetic information in life insurance underwriting from October 8^th^ 2026. A move which brings Australia, somewhat belatedly, into line with the United States and some European countries. In terms of other concerns raised by our patients, anxiety related to decision-making and uncertainty about results also emerged, highlighting the psychological dimensions of genetic testing. Additionally, equity of access and cost were noted, reflecting systemic challenges documented in prior cancer research. For example, a multinational study of 1,176 breast cancer patients across nine countries ^38^ concluded that accessible reimbursement policies for cancer genetic testing are urgently needed worldwide to alleviate the financial burden on patients and their families. While some countries do have policies in place, it was clear from this study by Powell and colleagues ^38^ that most patients in the surveyed countries had poor awareness of the eligibility criteria even if they did exist. Thus, providing patients with adequate and accessible information regarding these concerns is essential to ensuring equitable implementation and uptake of germline genetic testing for PrCa.

Patients articulated clear informational needs, including details on testing procedures, clinical utility, cost and data privacy. The preference for information delivery by trusted clinicians, particularly medical practitioners and genetic counsellors, reinforces the importance of integrating genetic counselling into routine care pathways. Other studies in the breast and ovarian cancer space have made similar recommendations, with a particular emphasis on engagement with a trusted provider ^39^ and a critical need for personalised support and counselling beyond the generic information, as a cancer diagnosis can be a very vulnerable time for patients and their families ^40^. Here, the suggestion of a PrCa nurse as an additional support reflects a desire for gender-specific and condition-specific expertise, which may improve patient comfort and engagement. Responses regarding the use of questionnaires in clinical practice indicated that most patients were comfortable sharing health-related information, though preferences varied between interviews and questionnaires. The recommendation for closed questions due to limited baseline understanding suggests that questionnaire design should balance simplicity with opportunities for open-ended responses where appropriate. Previous studies have found that brief and timely questionnaires were associated with the highest level of patient satisfaction ^41^, and questionnaires that captured emotional impact, understanding of results and family implications were most valued in a familial hypercholesterolemia study ^42^.

In *Phase II*, while approximately half of healthcare providers and most carers/family members perceived the toolkit’s purpose as clear, only one-third of patients shared this view. Patient identified areas for improvement included that the toolkit did not consistently communicate its intent in a way that resonated with patients. The fact that most carers/family members thought the toolkit was clear, highlights that carers and family members often engage with health information with a practical mindset, in which they evaluate the toolkit on the information provided rather than on personal resonance or emotional relevance ^43,44^. Our toolkit was designed to provide an understanding of the purpose and procedures of genetic testing, however our patient cohort indicated their desire for additional information pertaining to practical realities patients face when making healthcare decisions. Patient relevance must not be assumed, and information must be framed around patients’ lived experiences, immediate concerns and personal meaning ^45^. The inclusion of real patient stories, representing a range of patient experiences, may further enhance patient-centred framing by contextualising complex information, supporting emotional engagement and helping patients see themselves reflected in the material. A growing body of evidence demonstrates that narrative-based health communication can improve comprehension and perceived relevance particularly in areas such as cancer and genomics ^46^. Previous research indicates that patient-facing resources are more effective when the content is explicitly tailored to the intended audience, or when toolkits incorporate clearly delineated sections designed for different audiences (i.e., patients, carers/family members, healthcare providers) ^47^. In our study, concerns about privacy and genetic and insurance discrimination demonstrate that patients’ responses to health information are shaped by fear, uncertainty and lack of trust, not just knowledge gaps, which is consistent with other findings ^48^. Toolkits should therefore clearly communicate legal protections (if any are in place in the relevant country), privacy safeguards, and available financial support to mitigate fear and uncertainty about genetic disclosures.

All three groups generally agreed that medical terms were well defined, however, confusion around terms such as ‘panel’ and ‘PARP inhibitors’ among patients/carers/family members indicates that technical jargon is an important barrier to consider. Of the few studies examining patients’ practical concerns on the costs and implications for insurance regarding PARP inhibitors, they all highlighted the urgent need for a better understanding of patient needs and perspectives for these complex gene-based therapies (reviewed in Mukherjee *et al.,* 2025 ^49^). Our study aimed to directly address this critical gap, and we provide novel evidence in this patient-centred space. Patients and carers/family members prioritised inclusion of concerns to do with cost, access, insurance and treatment implications, whereas healthcare providers focussed more on missing procedural and technical content. While our toolkit in its current form is insufficient, our study provides valuable insights into patients’ understanding and perceptions of precision medicine and germline genetic testing for PrCa, providing a valuable foundation for a future toolkit (or next iteration) that supports informed decision-making.

Several limitations must also be considered when interpreting our study results. Our recruitment strategy, through knowledgeable and highly motivated community groups, may have unintentionally excluded individuals with a lower health literacy of cancer. Also, this strategy may not have captured the full range of patient experiences, although three of our patients had undergone genetic testing associated with their PrCa diagnosis. Furthermore, by collecting minimal clinical data, we were able to minimise participant burden; however, this approach also limited the extent to which we could characterise our patient cohort in detail. We also make note of our sample size but acknowledge that this is comparable with other focus group studies. In fact, a 2016 study found that 90% of core themes emerged within three focus groups and small group sizes of 3-6 participants were often sufficient ^25,50^. The group setting may have influenced how some patients responded, although an online survey was provided to patients, none of whom undertook this option. However, the group environment was also observed to have known benefits, enabling interaction and discussion among participants. As for *Phase II*, of our 32 participants, only four carers/family members participated. Whilst we must acknowledge this small sample size, we note that their opinions and perspectives are valued, and it enabled us to identify views not expressed in other groups. In the future, we will plan to include men who have had a recent diagnosis or who are unaffected but have a strong family history of PrCa. In addition, our data showed a divergence in perceived usefulness of the toolkit, which suggests that a single toolkit may not meet the needs of all groups equally.

In conclusion, our study used patient engagement to inform and co-develop a toolkit designed to support patients undergoing germline genetic testing for PrCa, the first of its kind. Significantly, our study addresses a critical gap between the growing clinical relevance of germline genetic testing for PrCa and the limited informational support currently available to help patients navigate this process. In the current clinical setting, integration of genetics into PrCa care pathways is complex and often difficult to navigate for both patients and healthcare providers. This patient-centred approach enhances the relevance, acceptability and usability of our toolkit, increasing its potential to support informed decision-making, reduce concerns and improve patient preparedness for germline genetic testing. In hindsight, a multi-stage approach allowing co-development with patients during the design of the toolkit would have been beneficial, prior to evaluation. Furthermore, future research to evaluate the toolkit’s impact on decision-making and uptake of genetic testing in real-world clinical settings, should include evaluation across treatment stages and cultural/socioeconomic contexts. As germline genetic testing becomes increasingly embedded in PrCa clinical pathways, consumer co-designed and tailored resources will be essential to ensuring that advances in genomic medicine lead to meaningful and inclusive improvements in patient outcomes.

## Data Availability

Additional data is included in the supplementary materials. All data produced in the present work is contained in the manuscript. A copy of the toolkit is available upon reasonable request to the authors.

## AUTHOR CONTRIBUTIONS

JLD, JR and LMF conceived the study. KR managed and led the study. KR, JR, RW and KB attended the focus groups. LB and JM performed the analyses. KR wrote the original draft and KR, LB and JM formatted the tables. All authors reviewed and edited the manuscript.

## ACKNOWLEDGEMENTS

We are greatly indebted to all our participants, particularly those men who participated in both phases of our study. We would also like to particularly thank our local, engaged healthcare providers, Professor Rosemary Harrup, A/Prof Marketa Skala, A/Prof Louise Nott and Dr Mathew Wallis, who helped secure funding, guide ethics and governance approvals and aided participant recruitment.

## DATA AND CODE AVAILABILITY

Additional data is included in the supplementary materials.

