## Supplementary for "What tools do men need to make an informed decision about germline genetic testing for prostate cancer? A qualitative and survey study"

### Supplementary Information

#### Supplementary Information 1. *Phase I* Focus Group Prompts

##### Focus Group Prompts – Indicative Questions to Facilitate Discussion

**Phase 1: Patients with Prostate Cancer**

*General information about your prostate cancer*

How old are you?

Could you please tell me a bit about your diagnosis of prostate cancer?

When were you first diagnosed with this condition?

Have you or are you having had any treatment for your prostate cancer since your diagnosis? What was/is that treatment?

Have you experienced side effects from current or previous treatments?

*Views on genetic testing /patient-reported outcome collection*

Sometimes people talk about the term 'precision medicine'. Have you heard about this? What does this mean to you? What do you think about it?

Have you heard of the use of genetic testing for the diagnosis and treatment of other cancers? What does this mean to you? What do you think about it?

If you were going to undertake genetic testing relating to your prostate cancer, what information would you hope it would provide about your disease?

What benefits do you think there might be? What would be your concerns?

Who would you like to talk to about genetic testing? What information do you think you would need? What support might you need?

In clinical care in cancer, people often want to obtain the patient's perspective on their experience, usually through questionnaires known as 'patient-reported outcome measures.' What aspects of a patient's experience with genetic testing would you want to talk about in questionnaires?

What aspects of a patient's experience with genetic testing would you want to talk about in questionnaires? Are there aspects you would not want to discuss or would not feel comfortable discussing?

#### Supplementary Information 2. *Phase II* Focus Group Prompts

##### Focus Group/Interview Prompts – Indicative Questions to Facilitate Discussion

###### Phase 2: Patients with Prostate Cancer

1. Do you think the purpose of the Information Toolkit is clear?
2. Do you understand how things are described in the toolkit?
3. When you see medical or scientific terms in the toolkit, do you think they are defined clearly? Do you understand them?
4. Are there medical/scientific or other terms in the toolkit that you don't understand?
5. Is the toolkit missing information that you would find help? What information would you like to add?
6. Is there any information that you found worrying or concerning?
7. Would you find this Information Toolkit helpful if you were offered genetic testing by your doctor or specialist?

*Additional questions for discussion (if time allows):*

8. The toolkit is broken into "chunks" of information in short sections. Do you find this helpful?
9. Do you think the headings of the short sections provide useful information?
10. Do you find the order of sections in the toolkit easy to follow? Does it make sense to you?
11. Would you prefer another way of ordering the toolkit sections?
12. Do the diagrams help you understand the content?
13. Do you find the photos in the toolkit distracting? Probe: Which ones do you like and dislike (1-2)?
14. Is the text big enough and easy to read?
15. Is the audio easy to understand? (e.g., not too fast, not garbled)
16. Do you feel like there is information that is repeated in the toolkit?
17. Do you have any other comments to add?

###### Phase 2: Prostate Cancer Healthcare Providers

1. Do you think the purpose of the Information Toolkit is clear?
2. Do you understand how things are described in the toolkit? Do you think your patients would understand how things are described in the toolkit?
3. When you see medical or scientific terms in the toolkit, do you think they are defined clearly? Do you think your patients would understand them?
4. Are there medical/scientific or other terms in the toolkit that you don't understand, or you think your patients wouldn't understand?

5. Is the toolkit missing information that you would find help? What information would you like to add?
6. Is there any information that you think would cause worry or concern for your patients? Or other healthcare providers?
7. Would you find this Information Toolkit helpful when explaining and offering genetic testing to your patients?

*Additional questions for discussion (if time allows):*

8. The toolkit is broken into "chunks" of information in short sections. Do you find this helpful?
9. Do you think the headings of the short sections provide useful information?
10. Do you find the order of sections in the toolkit easy to follow? Does it make sense to you?
11. Would you prefer another way of ordering the toolkit sections?
12. Do the diagrams help you understand the content?
13. Do you find the photos in the toolkit distracting? Probe: Which ones do you like and dislike (1-2)?
14. Is the text big enough and easy to read?
15. Is the audio easy to understand? (e.g., not too fast, not garbled)
16. Do you feel like there is information that is repeated in the toolkit?
17. Do you have any other comments to add?

**Supplementary Tables**

**Supplementary Table 1.** ***Phase I*** **focus group themes.**

| Name/Description | Files | References |
|---|---|---|
| **1. Precision medicine** | **5** | **32** |
| 1.1 Understanding | 5 | 26 |
| a) Avoiding unnecessary treatment and side effects | 1 | 1 |
| b) Don't understand | 4 | 9 |
| c) More effective treatment | 1 | 1 |
| d) More precise or targeted medicine | 2 | 4 |
| e) Targeted to cancer | 1 | 1 |
| f) Targeted to individual | 3 | 7 |
| g) Therapy focused on one location | 2 | 3 |
| 1.2 Attitudes | 2 | 6 |
| a) Equity of access and cost | 1 | 1 |
| b) Pro | 2 | 5 |
| c) Pro - if outcome is better | 1 | 1 |
| d) Pro - innovation is good | 1 | 2 |
| **2. Genetic testing** | **5** | **191** |
| 2.1 Understanding | 4 | 20 |
| a) Know familial risk | 3 | 6 |
| b) Mutations associated with cancer | 1 | 2 |
| c) Unsure or need more information | 4 | 8 |
| d) Currently no treatment following on | 1 | 1 |
| e) Unsure about additional benefits compared with current testing | 1 | 1 |
| f) Would like to know how genes are associated | 1 | 2 |
| g) Way of finding best treatment | 3 | 4 |
| 2.2 Attitudes | 4 | 9 |
| a) No complaints about usual care treatment (without testing) | 1 | 1 |
| b) Pro | 2 | 3 |
| c) Understanding familial risk would be helpful | 3 | 5 |
| 2.3 Information regarding disease | 4 | 24 |
| a) Implications for health insurance | 1 | 1 |
| b) Risk to family members | 4 | 9 |
| c) Risk to self | 3 | 6 |
| d) Information re cause - genetic or environmental | 1 | 1 |
| e) Level of aggressiveness | 2 | 2 |
| f) What to look out for | 1 | 1 |
| g) Treatment options with best outcomes | 3 | 8 |
| h) Potential for immunotherapy | 2 | 2 |
| 2.4 Benefits | 4 | 28 |
| a) Early detection and prevention | 2 | 5 |

| | | |
|---|---|---|
| b) For patient triage or targeting | 2 | 2 |
| c) Identify people at risk | 1 | 1 |
| d) Target resources by excluding patients | 1 | 1 |
| e) Help medical science | 1 | 1 |
| f) Identify best treatments | 3 | 6 |
| g) Inform family members of their risk | 3 | 7 |
| h) For family early detection | 2 | 3 |
| i) For family fertility decisions | 1 | 1 |
| j) Know what you're dealing with | 1 | 1 |
| k) More accurate than current testing | 1 | 1 |
| l) Outweigh risks | 1 | 1 |
| m) Reduce use of less effective or more onerous therapies or tests | 1 | 3 |
| n) Understand reasons for other diagnoses | 1 | 1 |
| 2.5 Concerns | 5 | 32 |
| a) Deciding whether it is worth knowing | 1 | 3 |
| b) Equity of access and cost | 2 | 3 |
| c) Implications for friends and family | 1 | 2 |
| d) Worry about communicating bad result to others | 1 | 1 |
| e) Worry for people around you | 1 | 1 |
| f) Implications for health insurance | 1 | 5 |
| g) Implications for other restrictions | 1 | 1 |
| h) Increased anxiety | 2 | 2 |
| i) Meaning of test poorly understood | 1 | 1 |
| j) No concerns | 3 | 8 |
| k) Privacy concerns | 1 | 2 |
| l) Side effect of surgery | 1 | 1 |
| m) Some doctors poor communicators | 1 | 1 |
| n) Worry about family members' risk | 2 | 3 |
| o) Worry about having children | 1 | 1 |
| 2.6 Desired information source | 4 | 20 |
| a) Genetic counsellor | 1 | 1 |
| b) Geneticist | 1 | 1 |
| c) Medical practitioners | 2 | 7 |
| d) GPs | 1 | 1 |
| e) Specialists | 1 | 1 |
| f) With special training | 1 | 3 |
| g) No counselling wanted | 1 | 2 |
| h) Non-specific person who knows what they're talking about | 1 | 2 |
| i) Researchers | 1 | 2 |
| 2.7 Desired information | 4 | 36 |
| a) Benefit | 1 | 2 |
| b) Benefit for choosing treatment | 1 | 1 |
| c) Best testing model | 1 | 1 |
| d) Cost | 2 | 2 |

| | | |
|---|---|---|
| e) Data risks | 1 | 2 |
| f) How information is used and collected | 1 | 1 |
| g) How reliable is the result | 1 | 1 |
| h) How to decide on treatment | 2 | 3 |
| i) If tested, can family members access result | 1 | 1 |
| j) Is the result indicator of risk or actual tumour | 1 | 1 |
| k) Method and experience of testing | 2 | 3 |
| l) No questions | 1 | 2 |
| m) Other considerations in deciding whether to test | 1 | 1 |
| n) Outcomes | 1 | 2 |
| o) Requires understanding of basic risk concepts | 2 | 2 |
| p) Risks - anxiety | 1 | 1 |
| q) Socioeconomic equity | 1 | 1 |
| r) Timing - for prevention or treatment guidance | 1 | 2 |
| s) Timing of results | 1 | 1 |
| t) What do the results mean | 1 | 1 |
| u) What information does the test give | 1 | 2 |
| v) What prompts testing | 1 | 2 |
| w) What questions to ask | 1 | 2 |
| 2.8 Desired support | 4 | 18 |
| a) Advice not just on clinical implications | 1 | 1 |
| b) Background information or counselling regarding what to do and ask | 1 | 1 |
| c) Male prostate cancer nurse | 2 | 2 |
| d) Multidisciplinary integrated team care | 1 | 6 |
| e) No support desired | 1 | 4 |
| f) Peer support | 1 | 1 |
| g) Psychology or counselling | 1 | 2 |
| h) Support person in appointments to take notes | 1 | 1 |
| **3. Questionnaires** | **5** | **23** |
| 3.1 Include | 2 | 2 |
| a) Do you have any questions | 1 | 1 |
| b) Environmental exposures | 1 | 1 |
| 3.2 Exclude | 4 | 7 |
| a) Nothing | 4 | 7 |
| b) Ensure simplicity | 1 | 2 |
| c) Lack of privacy as a cancer patient | 3 | 5 |
| d) Method of administration | 1 | 3 |
| e) Intimate information can be easier in written form | 1 | 1 |
| f) Need right environment to talk | 1 | 1 |
| g) Prefer interview to questionnaire | 1 | 1 |
| h) Questionnaire burden early in diagnosis | 1 | 1 |
| i) Take into account low understanding - closed rather than open questions | 1 | 2 |
| j) Would need to be confidential | 1 | 1 |
